# Inconsistent directions of change in case severity across successive SARS-CoV-2 variant waves suggests an unpredictable future

**DOI:** 10.1101/2022.03.24.22272915

**Authors:** David J. Pascall, Elen Vink, Rachel Blacow, Naomi Bulteel, Alasdair Campbell, Robyn Campbell, Sarah Clifford, Chris Davis, Ana da Silva Filipe, Noha El Sakka, Ludmila Fjodorova, Ruth Forrest, Emily Goldstein, Rory Gunson, John Haughney, Matthew T.G. Holden, Patrick Honour, Joseph Hughes, Edward James, Tim Lewis, Oscar MacLean, Martin McHugh, Guy Mollett, Tommy Nyberg, Yusuke Onishi, Ben Parcell, Surajit Ray, David L. Robertson, Shaun R. Seaman, Sharif Shabaan, James G. Shepherd, Katherine Smollett, Kate Templeton, Elizabeth Wastnedge, Craig Wilkie, Thomas Williams, The COVID-19 Genomics UK (COG-UK) consortium, Emma C. Thomson

## Abstract

**Objective:** To determine how the severity of successively dominant SARS-CoV-2 variants changed over the course of the COVID-19 pandemic.

**Design:** Retrospective cohort analysis.

**Setting:** Community- and hospital-sequenced COVID-19 cases in the NHS Greater Glasgow and Clyde (NHS GG&C) Health Board.

**Participants:** All sequenced non-nosocomial adult COVID-19 cases in NHS GG&C infected with the relevant SARS-CoV-2 lineages during analysis periods. B.1.177/Alpha: 1st November 2020 - 30th January 2021 (n = 1640). Alpha/Delta: 1st April - 30th June 2021 (n = 5552). AY.4.2 Delta/non-AY.4.2 Delta: 1st July - 31st October 2021 (n = 9613). Non-AY.4.2 Delta/Omicron: 1st - 31st December 2021 (n = 3858).

**Main outcome measures:** Admission to hospital, ICU, or death within 28 days of positive COVID-19 test

**Results:** For B.1.177/Alpha, 300 of 807 B.1.177 cases were recorded as hospitalised or worse, compared to 232 of 833 Alpha cases. After adjustment, the cumulative odds ratio was 1.51 (95% CI: 1.08-2.11) for Alpha versus B.1.177. For Alpha/Delta, 113 of 2104 Alpha cases were recorded as hospitalised or worse, compared to 230 of 3448 Delta cases. After adjustment, the cumulative odds ratio was 2.09 (95% CI: 1.42-3.08) for Delta versus Alpha. For non-AY.4.2 Delta/AY.4.2 Delta, 845 of 8644 non-AY.4.2 Delta cases were recorded as hospitalised or worse, compared to 101 of 969 AY.4.2 Delta cases. After adjustment, the cumulative odds ratio was 0.99 (95% CI: 0.76-1.27) for AY.4.2 Delta versus non-AY.4.2 Delta. For non-AY.4.2 Delta/Omicron, 30 of 1164 non-AY.4.2 Delta cases were recorded as hospitalised or worse, compared to 26 of 2694 Omicron cases. After adjustment, the median cumulative odds ratio was 0.49 (95% CI: 0.22-1.06) for Omicron versus non-AY.4.2 Delta.

**Conclusions:** The direction of change in disease severity between successively emerging SARS-CoV-2 variants of concern was inconsistent. This heterogeneity demonstrates that severity associated with future SARS-CoV-2 variants is unpredictable.

## Introduction

Since the SARS-CoV-2 pandemic started in late 2019, a succession of variants have achieved dominance, each replacing the previous dominant variant. From late 2020 these were designated variants of concern (VOCs); variants that exhibit increased transmission rates, antigenic differences, and/or case severity[1]. The three VOCs that most impacted both the pandemic and epidemic in Scotland were Alpha (Pango lineage B.1.1.7), which emerged in the UK in September 2020[2], Delta (Pango lineage B.1.617.2), which emerged in India prior to October 2020[1] and spread globally in May 2021, with >1000 introductions to the UK[3], and most recently Omicron (Pango lineage B.1.1.529), which emerged in Africa in November 2021[1], and spread very rapidly around the globe. Before the Omicron variant emerged, the Delta sublineage AY.4.2 was on course to replace the other Delta lineages, with growth rate estimates[4] implying that it would become dominant in the UK in early 2022. This spread was arrested by the emergence of the more transmissible and immune-evading Omicron variant, which has supplanted nearly all non-Omicron diversity[5].

Understanding any change in disease severity associated with infection by new variants of a virus (especially one that has newly entered the human population) is critical from a clinical, public health, and basic science perspective. For example, knowledge of the severity of a new variant is a vitally important in the decision-making process for the stringency of control measures and the roll out of vaccination and other treatments. It is expected that upon entry to a new host species from a zoonotic reservoir, the consequences of infection will be unpredictable with the severity of disease caused by the pathogen likely to be far from its evolutionary optima[6]. SARS-CoV-2 is on average associated with low virulence in younger age groups whilst severe outcomes manifest in older age groups and those with comorbidities. Evolution of virulence may therefore not be strongly constrained, rather, factors governing transmission and immune evasion are likely to determine the direction of evolution[7].

Our aim was to determine how the severity of successively dominant SARS-CoV-2 variants has changed over the course of the COVID-19 pandemic. To explore this, we prospectively linked detailed clinical metadata and viral genomic data from NHS GG&C to analyse relative case severity within 28 days of diagnosis between successive dominant lineages (Fig 1); B.1.177 versus Alpha, Alpha versus Delta, non-AY.4.2 versus AY.4.2 Delta, and non-AY.4.2 Delta versus Omicron. We test the robustness of these estimates to epidemic phase bias, a bias caused by patient outcomes being correlated with the time from infection to positive test, resulting in an estimated odds ratio adjusting for time of positive to test being a biased estimator of the odds ratio adjusting for time of infection[8]. When lineages differ in their incidence, this effect can lead to incorrectly concluding that one lineage is associated with more severe disease than the other, when the estimated difference is driven almost entirely by the epidemic phase bias. This series of comparisons allowed us to assess trends in severity, removing this bias and accounting for other critical variables including detailed comorbidity and changes in the availability of new treatments and vaccination.

**Figure 1.**
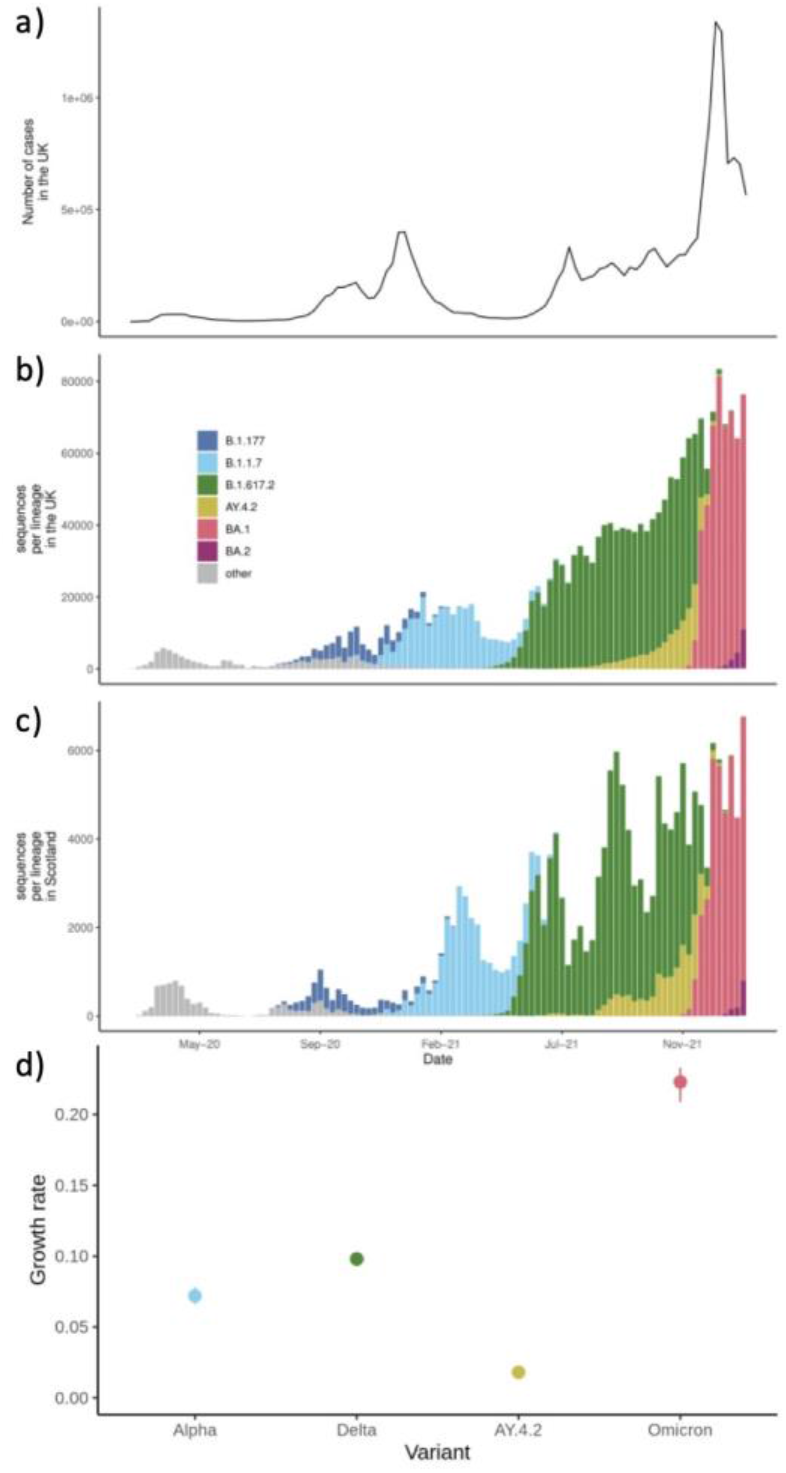
The history of the SARS-CoV-2 epidemic through time in the UK. A) Case UK case count over the course of the pandemic B) Number of sequences generated from the focal variants in the UK over the course of the pandemic C) Number of sequences generated from the focal variants in Scotland over the course of the pandemic D) The growth rates of each of the lineages over the period of their comparison

## Methods

This is a cohort study comparing the clinical outcomes of adults infected with a resident dominant SARS-CoV-2 lineage with those infected with the SARS-CoV-2 lineage that would displace it.

### Genome sequencing

Sequences were generated using the ARTIC Network protocol, originally developed for Oxford Nanopore-based sequencing[9]. and derived versions adapted to Illumina and ARTIC-unrelated amplicon-based protocols. The COG-UK pipeline was used for alignment and Pango lineage assignment[10].

### Data Inclusion/Exclusion

For the B.1.177/Alpha analysis, we included all sequenced samples with full data available on all adjustment variables from within the NHS GG&C health board between 1st November 2020 and 30th January 2021 (B.1.177: n = 807; Alpha: n = 833). A full demographic breakdown of samples is shown in Table S1. All sequences assigned as B.1.177 and associated sublineages were merged into a single category for the analysis. For the Alpha/Delta analysis we included all sequenced samples with full metadata between 1st April 2021 and 30th June 2021 (Alpha: n = 2104; Delta: n = 3448). All sequences assigned into B.1.617.2 and associated sublineages were merged into the Delta category for the analysis. A full demographic breakdown of samples is shown in Table S2. For the Delta/AY.4.2 analysis we included all sequenced samples with full metadata between 1st July 2021 and 31st October 2021 (non-AY.4.2 Delta: n = 8644; AY.4.2 Delta: n = 969). The AY.4.2 category was defined as all sequences assigned as either AY.4.2 or sublineages thereof, all remaining Delta sublineages were combined into the comparison category. A full demographic breakdown of samples is shown in Table S3. For the non-AY.4.2 Delta/Omicron analysis, we included all sequenced samples with full metadata between 1st December 2021 and 31st December 2021 (non-AY.4.2 Delta: 1164; Omicron: 2694). The Omicron category was defined as all sequences assigned as BA.1 or B.1.1.529, the Delta category was defined as for the Delta/AY.4.2 comparison. Full demographic breakdown of samples is shown in Table S4. Cases with hospital acquired COVID-19, defined as a first positive PCR test occurring more than 48 hours following admission to hospital, and all cases younger than 18 were excluded.

### Data lineage

Cohorts and de-identified linked data for the entire NHS GG&C health board (1.2 million people) were prepared by the West of Scotland Safe Haven at NHS GG&C. Data used in the analysis included maximum clinical severity at 28 days after the first positive test via a 4-point ordinal scale (1. No hospitalisation; 2. Hospitalisation (excluding elective surgery); 3. Admission to HDU/ICU; 4. Death), age at diagnosis, date of positive test, sex, partial postcode, number of vaccine doses, number of relevant comorbidities or risks of ill health (chronic cardiac disease, chronic respiratory disease, chronic renal disease, liver disease, dementia, chronic neurological conditions, connective tissue disease, diabetes, HIV infection, malignant tumours, clinician defined obesity, case shielding, immunosuppressive drugs, chemotherapy) and reinfection. Vaccination, comorbidities, reinfection, age and sex were included as they are known to impact clinical outcomes. Date of positive test was included to attempt to adjust for any time varying effects across the study window (e.g., changing clinical practice, hospital load, etc.). Partial postcode was included to attempt to account for spatial clustering of risk mediated though unmeasured variables (e.g., deprivation, ethnicity, etc.).

Severity was scored twice. For severity for “with” analyses, cases were assigned the most severe event that occurred within 28 days of their positive test. For the “of” analyses, events were only counted when they were explicitly linked to COVID-19 infection in the electronic patient records.Different datasets used in the linkage recorded “cause of” event differently. In datasets where ICD-10 codes were used (SMR01, accident and emergency, and deaths), a COVID-19 related ICD-10 code was required (specifically, any code starting U07, U04.9 which corresponds to an incorrect usage of the SARS ICD-10 code, and U10). In datasets where ICD-10 codes were not used (Scottish Intensive Care Society Audit Group data), the string “covid” was searched for case-insensitively in the free text entry. When the number of vaccine doses an individual had been given was calculated, if the last dose had been received less than 14 days before the date of the positive PCR test, it was ignored. Individuals with multiple confirmed episodes of infection (defined as separate positive PCR results more than 90 days apart) were marked as reinfected for any episodes after the first.

### Statistical analysis of clinical data

The four-level patient outcome data were analysed using cumulative generalised additive mixed models (GAMMs) with logit links[11] fit using Bayesian inference. These GAMMs included lineage, reinfection, patient sex and number of vaccine doses as categorical fixed effects and number of ISARIC4C identified comorbidities as a continuous fixed effect, with partial postcode included as a random effect. We included age and date of positive test as non-linear penalised regression splines. The basis dimension of the penalised regression splines was set to the number of unique dates of positive tests minus one and the number of unique ages (rounded to year) minus one respectively, with the intention that regularisation occur through the prior. Given that the pandemic was in its early stages during the first comparison (B.1.177/Alpha) and the vaccination campaign had not yet started, both reinfection and number vaccine doses received were excluded from the first model.

The same classes of parameter received the same priors in each model. The intercepts of the models were given t-distribution (location = 0, scale = 2.5, df = 3) priors, fixed effects were given normal (mean = 0, standard deviation = 2.5) priors, random effects and spline standard deviations were given exponential (mean = 2.5) priors. All severity models were fitted using the brms (v. 2.14.4) R package[12]. All presented models had no divergent transitions and effective sample sizes of over 200 for all parameters.

Sensitivity to epidemic phase bias was assessed using the method of Seaman *et al*.[8]. We added four days to the population who experienced more extreme outcomes (i.e., hospitalisation, admission to ICU/HCU or death), generating a modified test time for each individual, where if the patient was not hospitalised, it was their original test time and otherwise it was this new test time. We refit the model using the modified times and using individuals whose modified times lie within the inclusion window. The resulting estimate of the cumulate odds ratio was then compared with the cumulate odds ratio estimated from the original model.

### Growth rate estimates

We took the Scottish sequences and looked at the count of non-reference nonsynonymous mutations found each day and used the nlstools package in R[13] to model the daily logistic growth rate of each lineage. We took defining mutations for each lineage to model the growth rate of the lineage. The defining mutations for the variants were chosen as those which were not present in the previously dominant lineage. The defining mutations were N501Y for Alpha, L452R for Delta, A222V for AY.4.2, and N501Y for Omicron. For AY.4.2 we calculated the growth rate of A222V relative to L452R to represent the advantage relative to basal delta rather than against the shifting-in-proportion blend of Delta and Alpha variants. The period for each growth rate estimate covered the same window as the clinical analysis. The growth rate was taken from the growth parameter in the following equation of the regression. We used counts on both sides of the equation to down weight days with limited data providing noisier proportions.

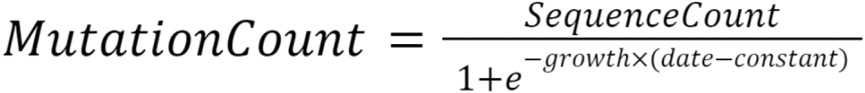

## Results

### B.1.177/Alpha

Our first comparison was the severity of the SARS-CoV-2 Alpha VOC versus the previous dominant non-VOC lineage (Pango designation B.1.177) (B.1.177: n = 807; Alpha: n = 833). The replacement of the lineages over time and their growth rates can be seen in Figure 1. We found that confirmed Alpha cases were associated with more severe infection (“of” - median cumulative odds ratio: 1.60; 95% central interval: 1.10-2.30 probability that effect is positive: >0.99; “with” – median cumulative odds ratio: 1.51; 95% central interval: 1.08-2.11; probability that effect is positive: 0.99; breakdown of number events in each group Table S1). The breakdown of “with” severity score by age and lineage can be seen in Fig 2a. Parameter estimates for all parameters can be found in Table S5. We found that, in our epidemic phase bias sensitivity analysis, Alpha remained associated with increased severity, but that the magnitude was reduced, as would be expected, and that in both analyses the probability of the effect being positive was reduced below 95% (“of” - median cumulative odds ratio: 1.31; 95% central interval: 0.91-1.88; probability that effect is positive: 0.93; “with” – median cumulative odds ratio: 1.24; 95% central interval: 0.88-1.79; probability that effect is positive: 0.88).

**Figure 2.**
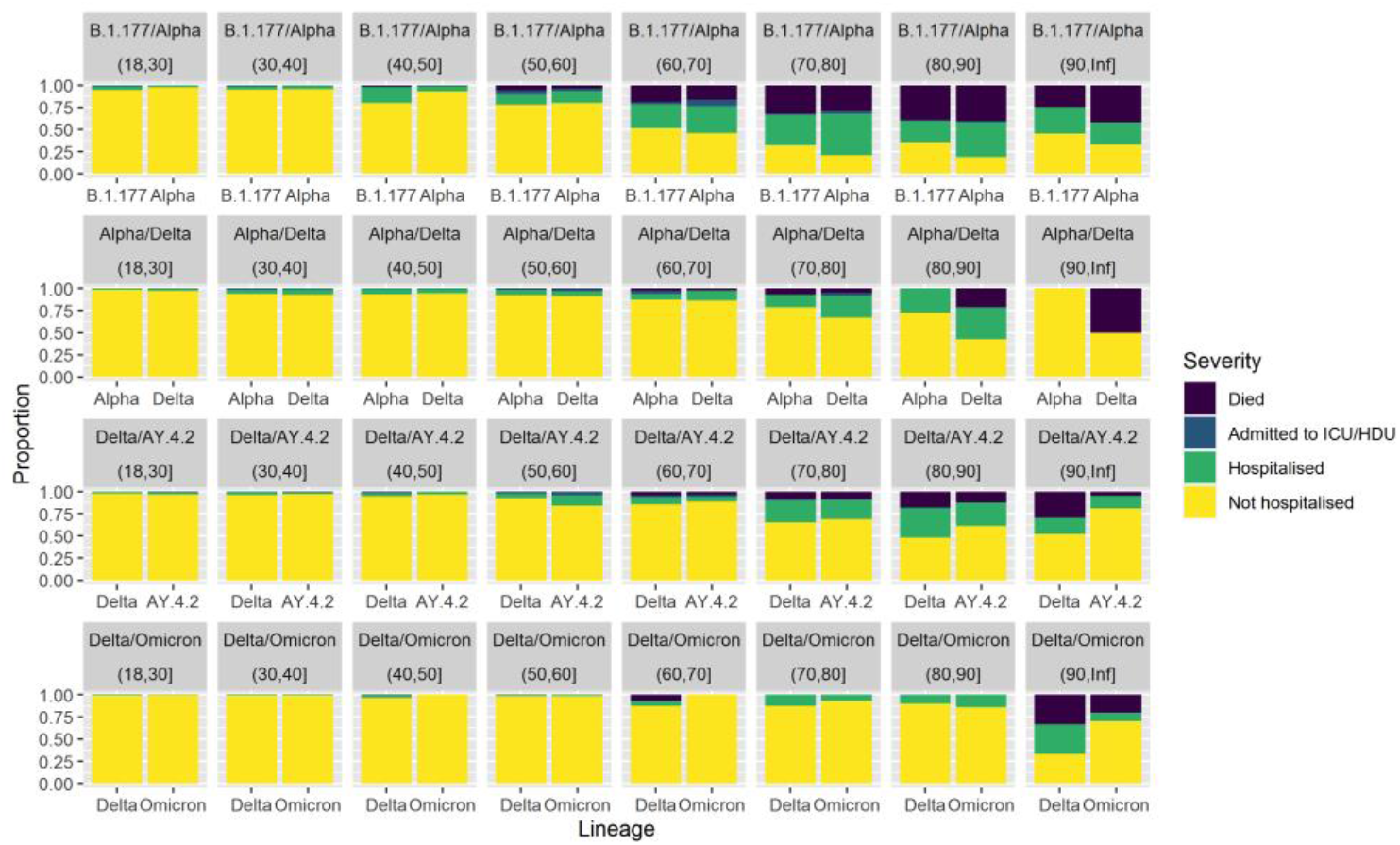
Comparison of case severity by age between pairs of co-circulating SARS-CoV-2 lineages. Clinical severity was measured on a four-level ordinal scale based on observed outcomes within 28 days of a positive test: no hospitalisation, hospitalisation, admission to ICE/HDU, death. The first row compares B.1.177 and Alpha between 1st November 2020 and 30th January 2021. The second row compares Alpha and Delta between 1st April 2021 and 30th June 2021. The third row compares Delta lineages against AY.4.2 between 1st July 2021 and 30th September 2021. The fourth row compares non-AY.4.2 Delta and Omicron between 1st December 2021 and 31st December 2021.

### Alpha/Delta

Our second comparison was the severity of the SARS-CoV-2 Delta VOC versus the previous dominant Alpha VOC lineage (Alpha: n = 2104; Delta: n = 3448). We estimate a substantial increase in case severity associated with Delta infections relative to Alpha (“of” - median cumulative odds ratio: 2.19; 95% central interval: 1.48-3.32; probability that effect is positive: >0.99; “with” - median cumulative odds ratio: 2.09; 95% central interval: 1.42-3.08; probability that effect is positive: >0.99; breakdown of number events in each group Table S2). The breakdown of “with” severity score by age and lineage can be seen in Fig 2b. Parameter estimates for all parameters can be found in Table S6. This effect is large enough that in epidemic phase bias sensitivity bias model, we still estimate a positive effect with a probability of positivity of over 0.95 (“of” - median cumulative odds ratio: 1.45; 95% central interval: 0.98-2.16; probability that effect is positive: 0.96; “with” - median cumulative odds ratio: 1.40; 95% central interval: 0.97-2.02; probability that effect is positive: 0.97).

### Non-AY.4.2 Delta/AY.4.2 Delta

Our third comparison was the severity of the SARS-CoV-2 Delta sublineage AY.4.2 versus the other Delta sublineages it was replacing (non-AY.4.2 Delta: n = 8644; AY.4.2 Delta: n = 969). We estimate that the AY.4.2 lineage infections are associated with similar case severity to that seen in other Delta sublineage infections (“of” - median cumulative odds ratio: 1.05; 95% central interval: 0.79-1.39; probability that effect is positive: 0.64; “with” - median cumulative odds ratio: 0.99; 95% central interval: 0.76-1.27; probability that effect is positive: 0.46; breakdown of number events in each group Table S3). The breakdown of “with” severity score by age and lineage can be seen in Fig 2c. Parameter estimates for all parameters can be found Table S7. In this case, there is no noticeable effect of epidemic phase bias, likely as the growth rate difference between the two variants was small (“of” - median cumulative odds ratio: 1.02; 95% central interval: 0.76-1.35; probability that effect is positive: 0.57; **“**with” - median cumulative odds ratio: 0.98; 95% central interval: 0.75-1.27; probability that effect is positive: 0.44).

### Non-AY.4.2 Delta/Omicron

Our final comparison was the severity of the SARS-CoV-2 Omicron VOC versus the non-AY.4.2 Delta sublineages (non-AY.4.2 Delta: 1164; Omicron: 2694). We find that Omicron (BA.1 sublineage) infection is associated with substantially less severe disease (“of” - median cumulative odds ratio: 0.15; 95% central interval: 0.01-1.48; probability that effect is positive: 0.06; “with” - median cumulative odds ratio: 0.49; 95% central interval: 0.22-1.06; probability that effect is positive: 0.04; breakdown of number events in each group Table S4). The breakdown of “with” severity score by age and lineage can be seen in Fig 2d. Parameter estimates for all parameters can be found in Table S8. Both the “with” and “of” analyses show a strong impact of epidemic phase bias. Omicron was the faster growing lineage, so the estimates of Omicron are driven more negative (“of” - median cumulative odds ratio: 0.07; 95% central interval: <0.01-0.68; probability that effect is positive: 0.01; “with” - median cumulative odds ratio: 0.19; 95% central interval: 0.08-0.42; probability that effect is positive: <0.01).

## Discussion

The principal findings of this study of the relative severity of COVID-19 cases caused by successive SARS-CoV-2 variant waves in Scotland was that Alpha was associated with more severe disease than B.1.177, Delta was associated with more severe disease than Alpha, non-AY.4.2 Delta and AY.4.2 Delta were associated with similar disease severity, and Omicron was associated with much less severe disease than non-AY.4.2 Delta. These conclusions were after accounting for comorbidities, changes in treatment, previous infection and vaccine availability, and robust to epidemic phase bias and the possibility of coincidental SARS-CoV-2 infection at admission to hospital. The successive replacements that we studied were not consistent in the direction of change in case severity.

Our study design has several strengths. It the first study to our knowledge to analyse the sequential replacement of variants throughout the pandemic with respect to the progression of severity attributable to virus evolution, and to use a consistent analytical approach across sequential SARS-CoV-2 lineages. Lauring et al 2022[14] is, in spirit, similar to our work, but their primary focus is not on the trajectory of severity, and they do not consider multiple time matched comparisons. Our approach takes a broad view of the definition of disease severity, by including community- and hospital-based cases and by considering a wider variety of clinical outcomes than those considered in most previous severity analyses. We also test the robustness of the severity analysis to epidemic phase bias and the impact of differences in the definition of severity outcomes (“of” analysis versus “with” analysis) across sequential variants. Our study does, however, have some limitations. Firstly, we only include cases with sequenced genomes, and thus our sample is biased towards cases with lower Ct, because these cases are more likely to have been sequenced. This is likely to be particularly important when the Ct distribution of infections differs between variants. The direction of the sampling bias on our results depends on which of the variants is associated with the lower Ct infections. Additionally, requiring sequences for our samples limits us to the set of individuals who have been tested by PCR, likely to represent hospitalised patients more than those in the community. Also, our sample size was not large enough to adjust for all the factors we would have liked to (e.g., fitting a dose by vaccine brand interaction, given that different brands are known to provide differential protection against different variants[7,15,16]).

There have been a series of other studies investigating each of the comparisons in our study individually. For Alpha versus previous variants, most previous studies have also estimated an increase in severity over extant diversity. A wide variety of end points have been used, 28-day mortality, hospitalisation, and an ordinal scale based around supplemental oxygen[17-27]. Our estimates are consistent with the majority of these studies. Our sample is smaller than was used in some of these studies, but we benefit from much higher resolution clinical data, and being able to control for comorbidities. When considering the Delta variant, two UK community analyses found that Delta (or an S-gene proxy) infections were associated with a higher risk of admission to hospital than with Alpha[28,29]. Comparable results were observed in Danish, US, and Canadian populations with a study in a Norwegian population being the exception[30-33]. The US and Canadian studies also found that Delta was associated with increased risk of ICU admission and death[31,32]. Most of these studies are therefore consistent with our results. Our estimate that AY.4.2 is associated with approximately the sample severity as other Delta sublineages is inconsistent with an analysis of the English population which found that confirmed AY.4.2 cases are associated with lower hospitalisation risk than cases associated with non-AY.4.2 Delta[34]. This inconsistency between our study and others may be explained by differences in the adjustment variables used, or because the larger sample size in Nyberg et al. 2022[33] allowed precise isolation of a small negative effect, with their effect estimate falling within our credible interval. Our results are consistent with studies from England, Scotland, Canada, and the US suggesting that Omicron infections are less severe than infections with Delta[14,35-37]. This reduction in case severity resulted in increased numbers of patients being admitted to hospital with a coincidental positive SARS-CoV-2 test rather than due to COVID-19, but this did not seem to overly impact our estimate of the severity of the variant relative to Delta.

It is important to appreciate measures of disease severity are highly context dependent. The clinical situation of the pandemic has shifted dramatically in Scotland during the study period, from a time with very little prior immunity to one with widespread vaccine and prior infection mediated immunity. Treatment availability with steroids, antivirals and antithrombotic agents has had a huge impact on reducing length of hospital stay and mortality[38-40]. Testing patterns have also changed dramatically across the study, with periods of higher and lower rates of testing. All these factors may impact the relative severity of variants. For this reason, our results cannot be used to compare the intrinsic case severity of variants that were not co-circulating at the same time.

Our results demonstrate that successive variants of SARS-CoV-2 are associated with inconsistent differences in disease severity after other factors are accounted for in the analysis, including comorbidities, vaccination, previous infection, and changes in treatment. In keeping with this finding, we have recently demonstrated fundamental changes in the life cycle of the Omicron variants BA.1 and BA.2 that may provide a biological explanation for the substantial drop in severity associated with this variant [7]. Omicron is associated with less cell-to-cell fusion and tropism for nasal epithelial cells rather than cells present in the lungs as well as an endosomal rather than a direct cell entry pathway. Given that the direction of the evolution of SARS-CoV-2 virulence has not been consistent over time and that the entry pathway of SARS-CoV-2 may be changed by single amino acid changes within the S2 domain of the spike protein [41], historical trends in severity cannot be used to predict the severity of future variants. However, once a variant has emerged, the likelihood of immune evasion and the method of cell entry may be estimated from the genome sequence. The relative reduction in severity seen with the Omicron variant should not make us complacent to the potential risks of future SARS-CoV-2 variants. Any increase in disease severity in a variant with similar transmissibility to Omicron could be devastating to health systems and communities. This study provides an important baseline for future research monitoring the relative severity of new variants and highlights the importance of ongoing genomic “early-warning” surveillance to detect new variants of concern in a timely manner.

## Data Availability

Raw data is personally identifiable, and so are not available, but aggregated data are provided in the supplementary materials.

## Acknowledgements

The authors would like to acknowledge the work of the West of Scotland Safe Haven team in supporting extractions and linkage to de-identified NHS patient datasets.

## Patient and Public Involvement

Patients or the public were not involved in the design, or conduct, or reporting, or dissemination plans of our research.

## Supplementary Appendix

**Table S1.** Breakdown of demographics for B.1.177/Alpha comparison. When one cell has a number of individuals less than 5, another cell is randomly chosen to be given imprecisely to preserve anonymity.

|  | B.1.177 | Alpha |
| --- | --- | --- |
| **“with” severity score** |  |  |
| Not hospitalised | 507 | 601 |
| Hospitalised | 154 | 134 |
| Admitted to HDU/ICU | 13 | 14 |
| Died | 133 | 84 |
| **“of” severity score** |  |  |
| Not hospitalised | 543 | 618 |
| Hospitalised | <180 | <160 |
| Admitted to HDU/ICU | <5 | <5 |
| Died | 86 | 59 |
| **Age** |  |  |
| 18-29 | 98 | 152 |
| 30-39 | 100 | 144 |
| 40-49 | 86 | 130 |
| 50-59 | 108 | 139 |
| 60-69 | 101 | 98 |
| 70-79 | 115 | 62 |
| 80-89 | 142 | 84 |
| 90+ | 57 | 24 |
| **Biological sex** |  |  |
| Female | 471 | 449 |
| Male | 336 | 384 |
| **Previous infection** |  |  |
| No | <810 | <840 |
| Yes | <5 | <5 |
| **Number of vaccine doses received** |  |  |
| 0 | 799 | <810 |
| 1 | <10 | 27 |
| 2 | <5 | <5 |
| **Number of ISARIC4C identified comorbidities** |  |  |
| 0 | 389 | 546 |
| 1 | 133 | 105 |
| 2 | 105 | 78 |
| 3 | 78 | <60 |
| 4 | 59 | 27 |
| 5 | <30 | 18 |
| 6 | 12 | <5 |
| 7 | <5 | <5 |
| 8 | <5 | <5 |

**Table S2.** Breakdown of demographics of Alpha/Delta comparison. When one cell has a number of individuals less than 5, another cell is randomly chosen to be given imprecisely to preserve anonymity.

|  | Alpha | Delta |
| --- | --- | --- |
| **“with” severity score** |  |  |
| Not hospitalised | 1991 | 3218 |
| Hospitalised | 90 | 186 |
| Admitted to HDU/ICU | 17 | 29 |
| Died | 6 | 15 |
| **“of” severity score** |  |  |
| Not hospitalised | 2004 | 3249 |
| Hospitalised | <100 | <190 |
| Admitted to HDU/ICU | <5 | <5 |
| Died | <5 | 11 |
| **Age** |  |  |
| 18-29 | 739 | <1250 |
| 30-39 | <580 | 887 |
| 40-49 | 382 | 633 |
| 50-59 | 247 | 426 |
| 60-69 | 120 | 167 |
| 70-79 | 28 | 61 |
| 80-89 | 11 | 28 |
| 90+ | <5 | <5 |
| **Biological sex** |  |  |
| Female | 1081 | 1683 |
| Male | 1023 | 1765 |
| **Previous infection** |  |  |
| No | 2085 | 3426 |
| Yes | 19 | 22 |
| **Number of vaccine doses received** |  |  |
| 0 | 1606 | 2022 |
| 1 | 438 | 1001 |
| 2 | 60 | 425 |
| **Number of ISARIC4C identified comorbidities** |  |  |
| 0 | 1817 | 2989 |
| 1 | <180 | 305 |
| 2 | 65 | 83 |
| 3 | 28 | <40 |
| 4 | 7 | 19 |
| 5 | 5 | 8 |
| 6 | <5 | <5 |
| 7 | <5 | <5 |
| 8 | <5 | <5 |

**Table S3.** Breakdown of demographics of non-AY.4.2 Delta/AY.4.2 Delta comparison. When one cell has a number of individuals less than 5, another cell is randomly chosen to be given imprecisely to preserve anonymity.

|  | non-AY.4.2 Delta | AY.4.2 Delta |
| --- | --- | --- |
| **“with” severity score** |  |  |
| Not hospitalised | 7799 | 868 |
| Hospitalised | 575 | 70 |
| Admitted to HDU/ICU | 76 | 12 |
| Died | 194 | 19 |
| **“of” severity score** |  |  |
| Not hospitalised | 7928 | 884 |
| Hospitalised | 580 | 69 |
| Admitted to HDU/ICU | 9 | <5 |
| Died | 127 | <20 |
| **Age** |  |  |
| 18-29 | 2217 | 211 |
| 30-39 | 1678 | 191 |
| 40-49 | 1472 | 163 |
| 50-59 | 1391 | 147 |
| 60-69 | 889 | 105 |
| 70-79 | 514 | 74 |
| 80-89 | 395 | 57 |
| 90+ | 88 | 21 |
| **Biological sex** |  |  |
| Female | 4641 | 513 |
| Male | 4003 | 456 |
| **Previous infection** |  |  |
| No | 8567 | 960 |
| Yes | 77 | 9 |
| **Number of vaccine doses received** |  |  |
| 0 | 2028 | 214 |
| 1 | 1433 | <110 |
| 2 | 5170 | 648 |
| 3 | 13 | <5 |
| **Number of ISARIC4C identified comorbidities** |  |  |
| 0 | 6597 | 709 |
| 1 | 1028 | 137 |
| 2 | 487 | 68 |
| 3 | <280 | 30 |
| 4 | 159 | <20 |
| 5 | 68 | 7 |
| 6 | 27 | <5 |
| 7 | 7 | <5 |
| 8 | <5 | <5 |

**Table S4.** Breakdown of demographics for non-AY.4.2 Delta/Omicron comparison. When one cell has a number of individuals less than 5, another cell is randomly chosen to be given imprecisely to preserve anonymity.

|  | non-AY.4.2 Delta | Omicron |
| --- | --- | --- |
| **“with” severity score** |  |  |
| Not hospitalised | 1134 | 2668 |
| Hospitalised | 22 | <30 |
| Admitted to HDU/ICU | <5 | <5 |
| Died | <10 | <5 |
| **“of” severity score** |  |  |
| Not hospitalised | <1170 | <2700 |
| Hospitalised | <5 | <5 |
| Admitted to HDU/ICU | <5 | <5 |
| Died | <5 | <5 |
| **Age** |  |  |
| 18-29 | 341 | 826 |
| 30-39 | 295 | 669 |
| 40-49 | 244 | 457 |
| 50-59 | 195 | 412 |
| 60-69 | 57 | 203 |
| 70-79 | 16 | 88 |
| 80-89 | 10 | 29 |
| 90 | 6 | 10 |
| **Biological sex** |  |  |
| Female | 621 | 1500 |
| Male | 543 | 1194 |
| **Previous infection** |  |  |
| No | 849 | 2471 |
| Yes | 315 | 223 |
| **Number of vaccine doses received** |  |  |
| 0 | 220 | 233 |
| 1 | 58 | 86 |
| 2 | 876 | 2350 |
| 3 | 10 | 25 |
| **Number of ISARIC4C identified comorbidities** |  |  |
| 0 | 980 | 2353 |
| 1 | 119 | 232 |
| 2 | 38 | <70 |
| 3 | <20 | 27 |
| 4 | 7 | 7 |
| 5 | <5 | 9 |
| 6 | 5 | <5 |
| 7 | <5 | <5 |

**Table S5.** Parameter estimates from B.1.177/Alpha model.

|  | “with” | “of” |
| --- | --- | --- |
| Alpha | 1.51 (1.08-2.11) | 1.60 (1.11-2.27) |
| Male sex | 1.37 (1.06-1.76) | 1.45 (1.11-1.87) |
| Comorbidities | 1.43 (1.32-1.55) | 1.48 (1.35-1.60) |

**Table S6.** Parameter estimates from Alpha/Delta model.

|  | “with” | “of” |
| --- | --- | --- |
| Delta | 2.09 (1.42-3.08) | 2.19 (1.48-3.32) |
| Male sex | 1.12 (0.89-1.42) | 1.12 (0.88-1.45) |
| Comorbidities | 2.25 (1.99-2.54) | 2.25 (1.98-2.57) |
| One vaccine dose | 0.18 (0.12-0.26) | 0.14 (0.09-0.22) |
| Two vaccine doses | 0.14 (0.08-0.23) | 0.12 (0.07-0.21) |
| Reinfected | 1.03 (0.22-3.61) | 0.29 (0.03-1.69) |

**Table S7.** Parameter estimates from non-AY.4.2 Delta/AY.4.2 Delta model.

|  | “with” | “of” |
| --- | --- | --- |
| AY.4.2 | 0.99 (0.76-1.27) | 1.05 (0.79-1.39) |
| Male sex | 1.20 (1.03-1.40) | 1.26 (1.07-1.51) |
| Comorbidities | 1.87 (1.77-1.99) | 1.93 (1.82-2.06) |
| One vaccine dose | 0.27 (0.19-0.38) | 0.24 (0.15-0.34) |
| Two vaccine doses | 0.12 (0.10-0.16) | 0.13 (0.10-0.17) |
| Three vaccine doses | 0.21 (0.06-0.63) | 0.20 (0.05-0.65) |
| Reinfected | 0.29 (0.07-0.93) | 0.36 (0.09-1.16) |

**Table S8.**
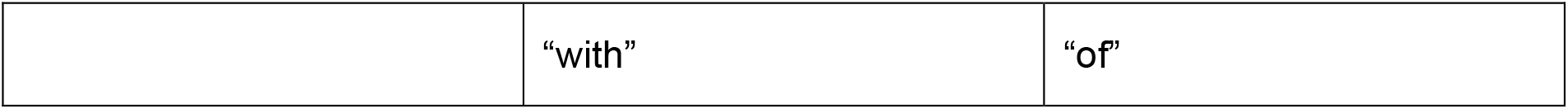
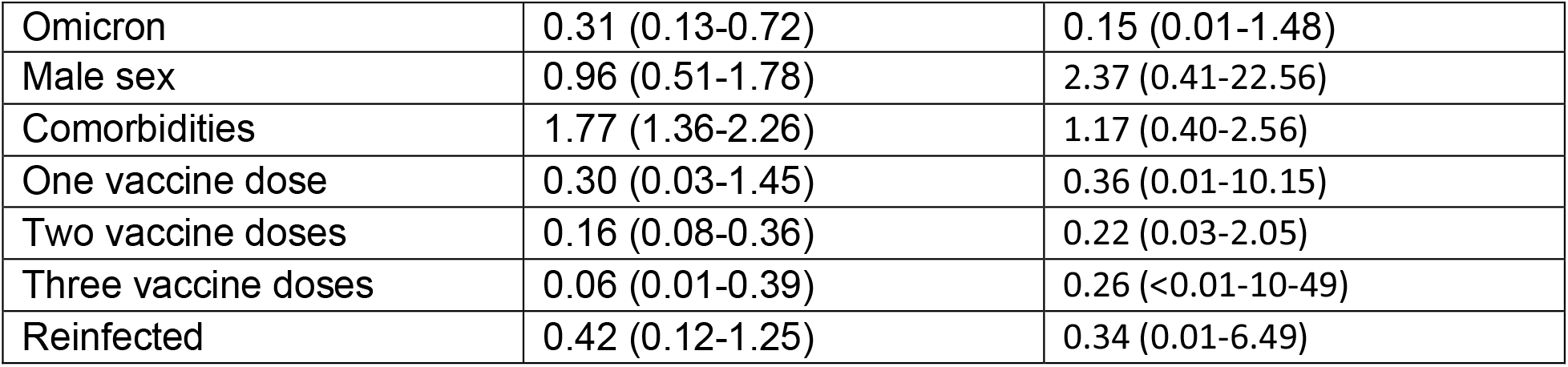
Parameter estimates from non-AY.4.2 Delta/Omicron model.

## References

1. World Health Organization [Internet]. Tracking SARS-CoV-2 variants. [cited 2022 March 23] Available from: https://www.who.int/en/activities/tracking-SARS-CoV-2-variants/

2. Rambaut A, Loman N, Pybus O, Barclay W, Barrett J, Carabelli A, et al. Preliminary genomic characterisation of an emergent SARS-CoV-2 lineage in the UK defined by a novel set of spike mutations. 2020 [cited 2022 March 23]. Available from: https://virological.org/t/preliminary-genomic-characterisation-of-an-emergent-sars-cov-2-lineage-in-the-uk-defined-by-a-novel-set-of-spike-mutations/563

3. McCrone JT, Hill V, Bajaj S, Pena RE, Lambert BC, Inward R, et al. Context-specific emergence and growth of the SARS-CoV-2 Delta variant. Nature. 2022. In press.

4. UK Health Security Agency. SARS-CoV-2 variants of concern and variants under investigation in England: Technical briefing 29. 2021 [cited 2022 March 23]. Available from: https://assets.publishing.service.gov.uk/government/uploads/system/uploads/attachment_data/file/1036501/Technical_Briefing_29_published_26_November_2021.pdf

5. GISAID [Internet]. Tracking of variants. [cited 2022 March 23]. Available from: https://www.gisaid.org/hcov19-variants/

6. Visher E, Evensen C, Guth S, Lai E, Norfolk M, Rozins C, et al. The three Ts of virulence evolution during zoonotic emergence. Proc Royal Soc B. 2021. 288:20210900.

7. Willett BJ, Grove J, MacLean OA, Wilkie C, De Lorenzo G, Furnon W, et al. SARS-CoV-2 Omicron is an immune escape variant with an altered cell entry pathway. Nat Microbiol. 2022. 7:1161–1179.

8. Seaman SR, Nyberg T, Overton CE, Pascall DJ, Presanis AM, De Angelis D. Adjusting for time of infection or positive test when estimating the risk of a post-infection outcome in an epidemic. Stat Methods Med Res. 2022. doi:10.1177/09622802221107105

9. Quick J. nCoV-2019 sequencing protocol v3 (LoCost) V.3. protocols.io. 2020 [cited 2022 March 23]. Available from: https://www.protocols.io/view/ncov-2019-sequencing-protocol-v3-locost-bp2l6n26rgqe/v3

10. Nicholls SM, Poplawski R, Bull MJ, Underwood A, Chapman M, Abu-Dahab K, et al. CLIMB-COVID: continuous integration supporting decentralised sequencing for SARS-CoV-2 genomic surveillance. BMC Genom. 2021. 22:196.

11. Bürkner P-C, Vuorre M. Ordinal Regression Models in Psychology: A Tutorial. Adv Meth Pract Psychol Sci. 2019. 2(1):77–101.

12. Bürkner P-C. brms: An R Package for Bayesian Multilevel Models Using Stan. J Stat Softw. 2017. 80(1):1–28.

13. Baty R, Ritz C, Charles S, Brutsche M, Flandrois J, Delignette-Muller M. A Toolbox for Nonlinear Regression in R: The Package nlstools. J Stat Softw. 2015. 66(5):1–21.

14. Lauring AS, Tenforde MW, Chappell JD, Gaglani M, Ginde AA, McNeal T. Clinical severity of, and effectiveness of mRNA vaccines against, covid-19 from omicron, delta, and alpha SARS-CoV-2 variants in the United States: prospective observational study. BMJ. 2022. 376:e069761.

15. Cele S, Jackson L, Khoury DS, Khan K, Moyo-Gwete T, Tegally H. Omicron extensively but incompletely escapes Pfizer BNT162b2 neutralization. Nature. 2021. 206:654–656.

16. UK Health Security Agency. SARS-CoV-2 variants of concern and variants under investigation in England: Technical briefing 34. 2022 [cited 2022 March 23]. Available from: https://assets.publishing.service.gov.uk/government/uploads/system/uploads/attachment_data/file/1050236/technical-briefing-34-14-january-2022.pdf

17. Davies NG, Jarvis CI, CMMID COVID-19 Working Group, Edmunds WJ, Jewell NP, Diaz-Ordaz K, et al. Increased mortality in community-tested cases of SARS-CoV-2 lineage B.1.1.7. Nature 2021. 593:270–274.

18. Challen R, Brooks-Pollock E, Read J, Dyson L, Tsaneva-Atanasova K, Danon L. Risk of mortality in patients infected with SARS-CoV-2 variant of concern 202012/1: matched cohort study. BMJ. 2021. 372:579.

19. Grint DJ, Wing K, Williamson E, McDonald HI, Evans D, Evans SJW, et al. Case fatality risk of the SARS-CoV-2 variant of concern B.1.1.7 in England, 16 November to 5 February. Euro Surveill. 2021. 26(11):2100256.

20. Nyberg T, Twohig KA, Harris RJ, Seaman SR, Flannagan J, Allen H, et al. Risk of hospital admission for patients with SARS-CoV-2 variant B.1.1.7: cohort analysis. BMJ. 2021. 373:1412.

21. Dabrera G, Allen H, Zaidi A, Twohig K, Thelwall S, Marchant E, et al. Assessment of Mortality and Hospital Admissions Associated with Confirmed Infection with SARS-CoV-2 Variant of Concern VOC-202012/01 (B.1.1.7) a Matched Cohort and Time-to-Event Analysis. SSRN. 2021. Available from: https://ssrn.com/abstract=3802578

22. Grint DJ, Wing K, Houlihan C, Gibbs HP, Evans SJW, Williamson E, et al. Severity of Severe Acute Respiratory System Coronavirus 2 (SARS-CoV-2) Alpha Variant (B.1.1.7) in England. Clin Infect Dis. 2021. ciab754.

23. Snell LB, Wang W, Alcolea-Medina A, Charalampous T, Batra R, de Jongh L, et al. Descriptive comparison of admission characteristics between pandemic waves and multivariable analysis of the association of the Alpha variant (B.1.1.7 lineage) of SARS-CoV-2 with disease severity in inner London. BMJ Open. 2022. 12:e055474.

24. Frampton D, Rampling T, Cross A, Bailey H, Heaney J, Byott M, et al. Genomic characteristics and clinical effect of the emergent SARS-CoV-2 B.1.1.7 lineage in London, UK: a whole-genome sequencing and hospital-based cohort study. Lancet Infect Dis. 2021. 21:1246–1256.

25. Ong SWX, Chiew CJ, Ang LW, Mak T-M, Cui L, Toh MPHS, et al. Clinical and Virological Features of Severe Acute Respiratory Syndrome Coronavirus 2 (SARS-CoV-2) Variants of Concern: A Retrospective Cohort Study Comparing B.1.1.7 (Alpha), B.1.351 (Beta), and B.1.617.2 (Delta). Clin Infect Dis. 2021. ciab721.

26. Stirrup O, Boshier F, Venturini C, Guerra-Assunção A, Alcolea-Medina A, Beckett A, et al. SARS-CoV-2 lineage B.1.1.7 is associated with greater disease severity among hospitalised women but not men: multicentre cohort study. BMJ Open Resp Res. 2021. 8:e001029.

27. Pascall DJ, Vink E, Blacow R, Bulteel N, Campbell A, Campbell R, et al. The SARS-CoV-2 Alpha variant is associated with increased clinical severity of disease in Scotland: a genomics-based prospective cohort analysis. medRxiv. 2022 [cited 2022 August 11]. Available from: https://doi.org/10.1101/2021.08.17.21260128

28. Sheikh A, McMenamin J, Taylor B, Robertson C, Public Health Scotland, EAVE II Collaborators. SARS-CoV-2 Delta VOC in Scotland: demographics, risk of hospital admission, and vaccine effectiveness. Lancet. 2021. 397(10293):2461–2462.

29. Twohig KA, Nyberg T, Zaidi A, Thelwall S, Sinnathamby MA, Aliabadi S, et al. Hospital admission and emergency care attendance risk for SARS-CoV-2 delta (B.1.617.2) compared with alpha (B.1.1.7) variants of concern: a cohort study. Lancet Infect Dis. 2022. 22(1):35–42.

30. Bager P, Wohlfahrt J, Rasmussen M, Albertsen M, Krause TG. Hospitalisation associated with SARS-CoV-2 delta variant in Denmark. Lancet Infect Dis. 2021. 21(10):1351.

31. Bast E, Tang F, Dahn F, Palacio A. Increased risk of hospitalisation and death with the delta variant in the USA. Lancet Infect Dis. 2021. 21(12):1629–1630.

32. Fisman DN, Tuite AR. Evaluation of the relative virulence of novel SARS-CoV-2 variants: a retrospective cohort study in Ontario, Canada. CMAJ. 2021. 193(42):E1619–E1625.

33. Veneti L, Salamanca BV, Seppälä E, Starrfelt J, Storm ML, Bragstad K, et al. No difference in risk of hospitalization between reported cases of the SARS-CoV-2 Delta variant and Alpha variant in Norway. Int J Infect Dis. 2022. 115:178–184.

34. Nyberg T, Harman K, Zaidi A, Seaman SR, Andrews N, Nash SG, et al. Hospitalization and Mortality Risk for COVID-19 Cases With SARS-CoV-2 AY.4.2 (VUI-21OCT-01) Compared to Non-AY.4.2 Delta Variant Sublineages. J Infect Dis. 2022. Jiac063.

35. Nyberg T, Ferguson NM, Nash SG, Webster HH, Flaxman S, Andrews N, et al. Comparative analysis of the risks of hospitalisation and death associated with SARS-CoV-2 omicron (B.1.1.529) and delta (B.1.617.2) variants in England: a cohort study. Lancet. 2022. In press.

36. Ulloa AC, Buchan SA, Daneman N, Brown KA. Estimates of SARS-CoV-2 Omicron Variant Severity in Ontario, Canada. JAMA. 2022. Online doi:10.1001/jama.2022.2274

37. Sheikh A, Kerr S, Woolhouse M, McMenamin J, Robertson C. Severity of omicron variant of concern and effectiveness of vaccine boosters against symptomatic disease in Scotland (EAVE II): a national cohort study with nested test-negative design. Lancet Infect Dis. 2022. 22:959–966.

38. The RECOVERY Collaborative Group. Dexamethasone in Hospitalized Patients with Covid-19. N Engl J Med. 2021. 384(8):693–704.

39. Weinreich DM, Sivapalasingam S, Norton T, Ali S, Gao H, Bhore R, et al. REGN-COV2, a Neutralizing Antibody Cocktail, in Outpatients with Covid-19. N Engl J Med. 2021. 384(3):238–251.

40. Sholzberg M, Tang GH, Rahhal H, Al Hamzah M, Kreuziger LB, Áinle FN, et al. Effectiveness of therapeutic heparin versus prophylactic heparin on death, mechanical ventilation, or intensive care unit admission in moderately ill patients with covid-19 admitted to hospital: RAPID randomised clinical trial. BMJ. 2021. 375:n2400.

41. Peacock TP, Brown JC, Zhou J, Thakur N, Sukhova K, Newman J, et al. The altered entry pathway and antigenic distance of the SARS-CoV-2 Omicron variant map to separate domains of spike protein. bioRxiv. 2022 [cited 2022 August 12]. Available from: https://doi.org/10.1101/2021.12.31.474653

